# Association of cortical arousals with sleep-disordered breathing events

**DOI:** 10.1101/2022.05.14.22275088

**Authors:** Kirsi-Marja Zitting, Brandon J Lockyer, Ali Azarbarzin, Scott A Sands, Wei Wang, Andrew Wellman, Stuart F Quan

## Abstract

**Study Objectives:** The American Academy of Sleep Medicine recommends scoring hypopneas in adults when there is a ≥3% oxygen desaturation Or when the event is associated with an arousal. However, there is no rule regarding the duration of the interval between the event termination and the associated arousal. The purpose of this study is to explore the timing between arousals and sleep-disordered breathing (SDB) events.

**Methods:** We analyzed cortical arousals (>1.6 million) and SDB events (>350,000 apneas and >1.9 million hypopneas) from 11,400 manually scored polysomnography recordings. Only arousals that started within ±30 seconds from the end of SDB events were included. We used the two local minimums on either side of the arousal distribution as the start/end times for the distribution, and to define which arousals are associated with SDB events. Finally, we calculated arousal probability near the end of SDB events.

**Results:** Cortical arousals with start times that fell within the two minimums were considered to be associated with SDB events. Using this definition, we found that 90% of apnea-associated arousals started no earlier than 4 seconds before and no later than 9 seconds after the end of apneas. Similarly, 90% of hypopnea-associated arousals started no earlier than 6 seconds before and no later than 14 seconds after the end of hypopneas, with the peak of the distribution coinciding with event end time. Arousal probability was highest during the first 10 seconds after the end of the event and was higher for longer events.

**Conclusions:** Our results suggest that 90% of SDB-associated arousals start from 6 seconds before to 14 seconds after the end of the respiratory events.

## INTRODUCTION

Cortical arousals are defined as abrupt shifts in the electroencephalogram (EEG) to a higher frequency (alpha and theta frequencies and frequencies >16 Hz) that last at least three seconds and are preceded by at least ten seconds of stable sleep.^1-4^ They occur frequently in association with the end of sleep-disordered breathing (SDB) events in sleep-related breathing disorders such as obstructive sleep apnea (OSA), although the mechanism by which respiratory stimuli induce arousals is still not clear.^5-8^ The current version (version 2.6) of the American Academy of Sleep Medicine (AASM) Manual for the Scoring of Sleep and Associated Events recommends scoring hypopneas in adults when there is a ≥ 3% oxygen desaturation from pre-event baseline or when the event is associated with an arousal, although it is also acceptable to score hypopneas in adults when there is a ≥ 4% oxygen desaturation from pre-event baseline,^1-3^ regardless of the presence of an arousal. The arousal criterion was added in 2012 due to the wide acceptance that SDB events associated with arousals cause significant and potentially dangerous sleep apnea symptoms, regardless of oxygen desaturation.^9^

However, the current rule provides little information on how to link cortical arousals to SDB events. Importantly, one needs to know the interval allowed between the end of a SDB event (either apnea or hypopnea) and the onset of the other event to determine if events are associated with each other. This deficiency may, in part, explain the large differences in apnea-hypopnea indices (AHIs) when the recommended and alternative rules are used for scoring hypopneas in adults.^10^

The purpose of this study is to explore the relationship between cortical arousals and SDB events (apneas and hypopneas), and to quantify the interval between arousals and the end of SDB events in a large number (11,400) of manually scored adult polysomnography recordings.

## METHODS

### Study Approval

This study uses de-identified data and does not meet the criteria for human subjects research as defined under federal regulations (45 CFR 46.102(f) of the Health and Human Services Policy for Protection of Human Research Subjects) and as confirmed by the Mass General Brigham Institutional Review Board (MGB-IRB), which reviewed the study protocol (2019P003560).

### Datasets

Ten well-characterized studies that had both sleep-wake and SDB event scoring available were selected for analysis (Table 1). The data for the three community cohort studies (SHHS, MESA, and SOF) as well as the data for one clinical research study (ABC) are publicly available at the National Sleep Research Resource (NSRR).^11^ The data for the other six other clinical research studies, for the ATS,^12^ the MPCS,^13^ the CTS,^14-17^ the CHFS,^18^ the MSS,^19^ and the TS^20^ studies, come from studies conducted in the Division of Sleep and Circadian Disorders at Brigham and Women’s Hospital (BWH). The Sleep Heart Health Study (SHHS)^21^ dataset includes data from the first (SSHS1) and second examinations of the cohort (SHHS2).

**Table 1.**
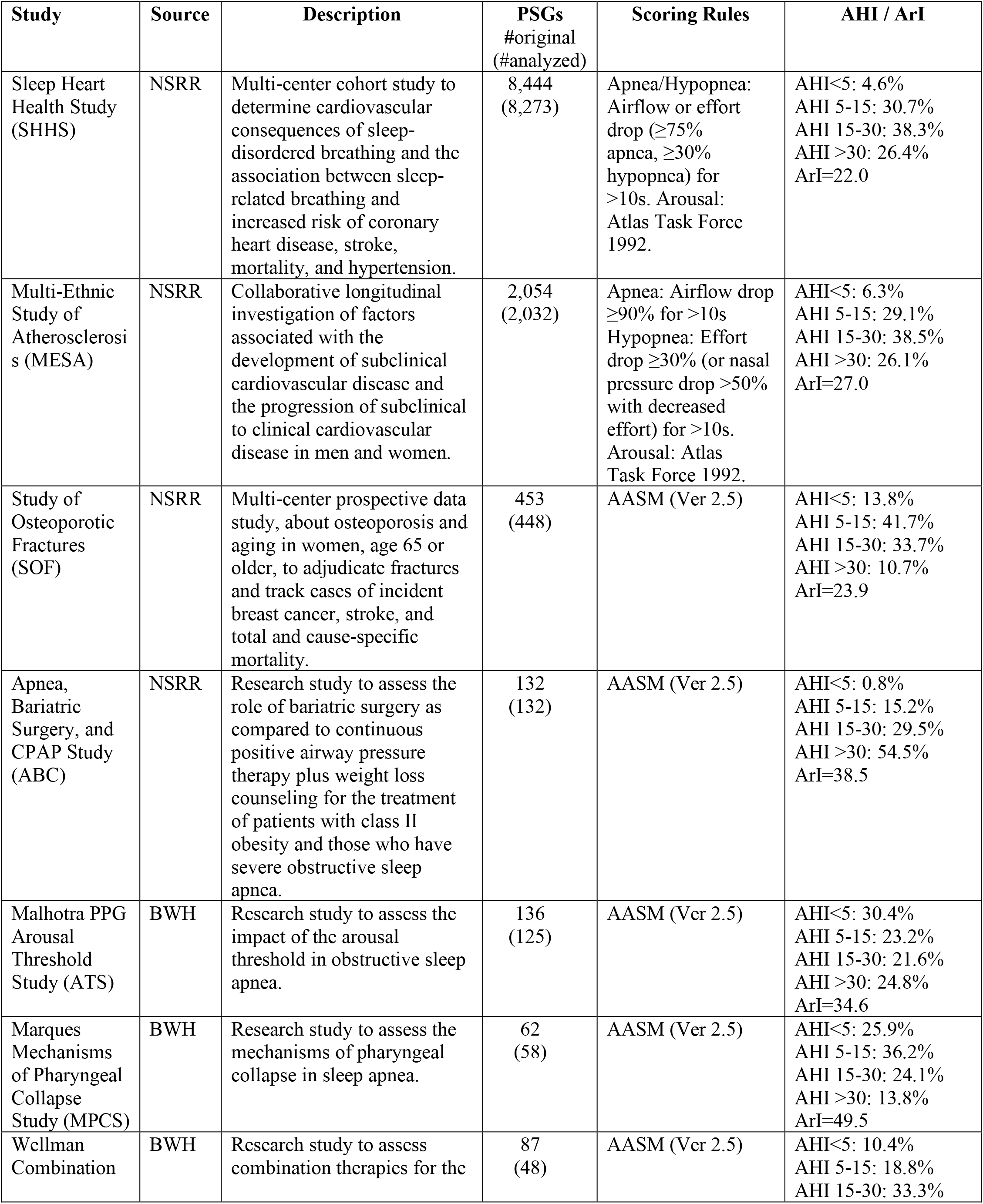
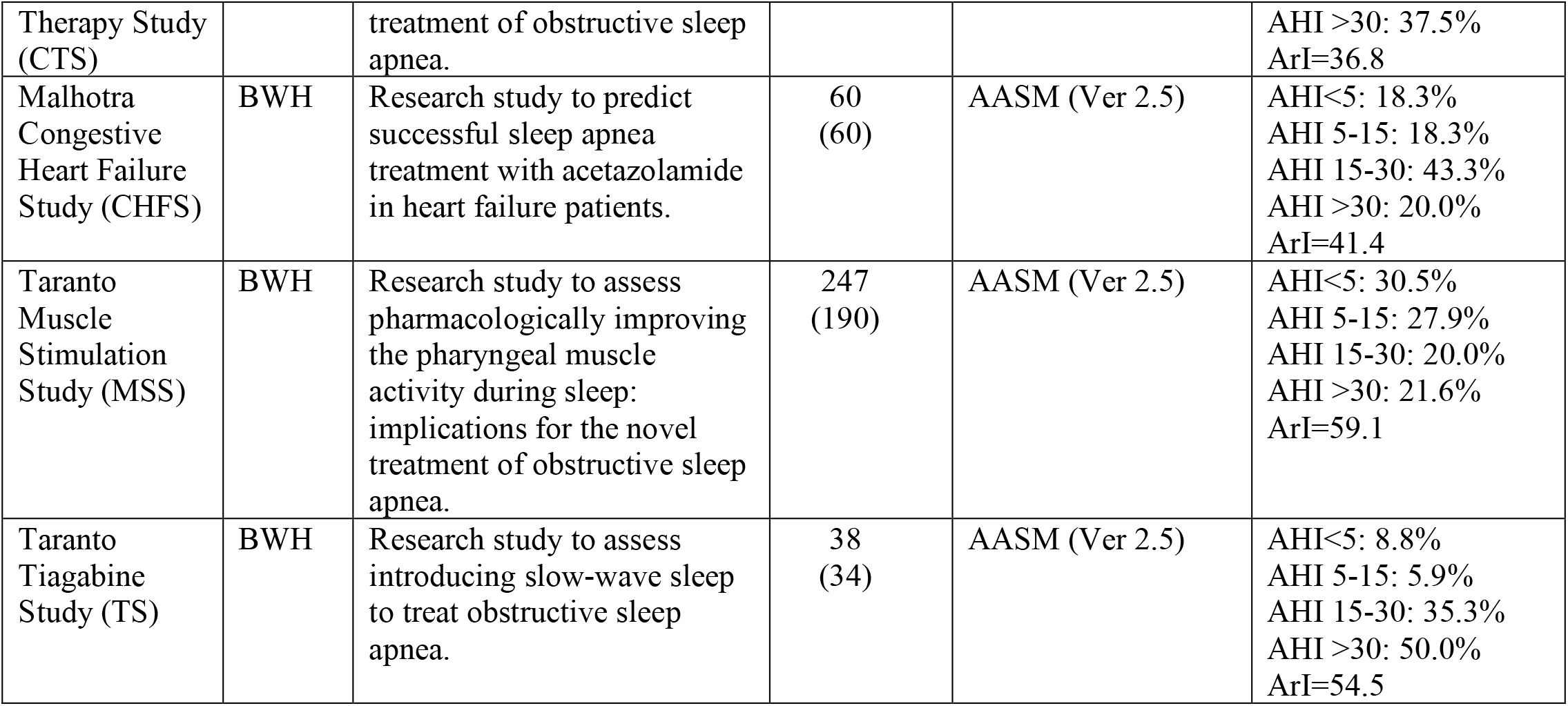
Datasets. National Sleep Research Resource (NSRR); Brigham and Women’s Hospital (BWH). The total number of recordings includes the original number of records (11,713) as well as the number of analyzed records (11,400) shown in parenthesis.

### Scoring Rules

The polysomnography (PSG) records had been scored according to established scoring rules (see Table 1). Briefly, the SHHS records were scored for apneas when airflow decreased ≥75% for ≥10 seconds, for hypopneas when any respiratory signal decreased ≥30% for ≥10 seconds (presence of desaturation and/or arousal not necessary), and for arousals according to the rules established in the Atlas Task Force, EEG Arousals: Scoring Rules and Examples (The Atlas Task Force, Sleep 1992). The MESA records were scored for apneas when airflow decreased ≥90% for ≥10 seconds, for hypopneas when respiratory effort decreased ≥30% for ≥10 seconds or nasal pressure decreased >50% for ≥10 seconds along with a decrease in respiratory effort (presence of desaturation and/or arousal not necessary), and for arousals according to the rules established in the Atlas Task Force, EEG Arousals: Scoring Rules and Examples (The Atlas Task Force, Sleep 1992). The SOF records were scored for apneas when there was a cessation of airflow for ≥10 seconds, for hypopneas when airflow decreased ≥30% for ≥10 seconds and was associated with a ≥3% desaturation or arousal, and for arousals according to the AASM criteria. The ABC records were scored for apneas when airflow decreased ≥90% for ≥10 seconds, for hypopneas when airflow decreased ≥30% for ≥10 seconds with a ≥4% desaturation, and for arousals according to the AASM criteria. The BWH records were scored for apneas when airflow decreased ≥90% for ≥10 seconds, for hypopneas when airflow decreased ≥30% for ≥10 seconds and was associated with a ≥3% desaturation or arousal, and for arousals according to the AASM criteria. In addition, the BWH records for the MCHF and the CTS studies also used ≥0% and ≥ 2% desaturation rules for hypopnea scoring when oxygen was administered.

### Data Processing

A total of 11,713 PSG records that had been manually scored for sleep-wake, SDB events, cortical arousals, and/or other events by various expert scorers according to established rules were available from these ten studies. The name of the recording and all event data (17,055,668 events) including event type, event start time and event duration, were extracted from the scoring output files in Python (version 3.8). Event labels for apneas, hypopneas, cortical arousals, and sleep stages were standardized across the datasets, and all central, mixed, and obstructive events were grouped into one category separately for apneas and hypopneas. Next, all duplicate events were removed, overlapping events of the same type were combined, the start and end time of the sleep period was identified from the first and last epochs of sleep, and total sleep time (TST) was calculated in minutes for each recording as follows: TST = N1 + N2 + N3 + R, where non-rapid eye movement sleep is indicated with the letter N, sleep stage within NREM is indicated with a number, and rapid-eye movement sleep is indicated with the letter R. Recordings with <1 hour of sleep and those with 0 arousals and/or 0 apneas or hypopneas were removed, and events that fell outside the sleep period were removed, which resulted in 11,400 recordings with 3,959,708 apnea, hypopnea, and arousal events (Figure 1). Apnea-hypopnea index (AHI) was calculated for each PSG record according to the AASM rule (version 2.6). Arousal index (ArI) was calculated for each record by dividing the total number of arousals by the number of hours of sleep.

**Figure 1.**
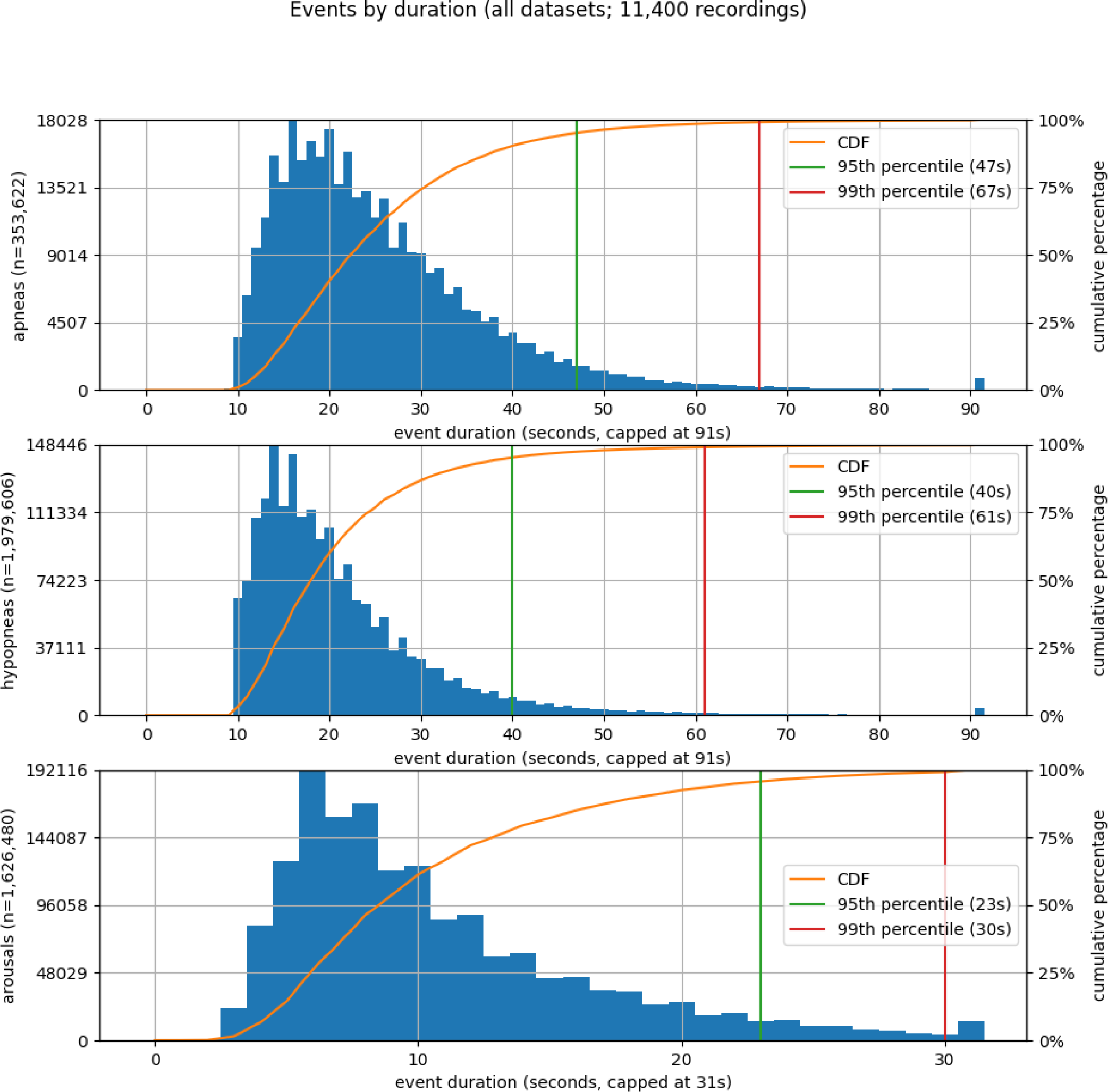
Frequency distribution histograms showing the total number (y-axis, left), cumulative percentage (y-axis, right), and duration (x-axis, in seconds) of apneas (upper panel), hypopneas (middle panel), and cortical arousals (lower panel) across all data (11,400 records). The x-axis shows event duration and ranges from 0 to 91 seconds for apneas and hypopneas, with events <10 seconds excluded and events >90 seconds included in the 91-second bin. The x-axis for cortical arousals ranges from 0 to 31 seconds for cortical arousals, with events <3 seconds excluded and events >30 seconds included in the 31-second bin. The orange line shows the cumulative distribution function (CDF) for the number of events, while the green and red lines indicate the maximum event durations within which 95% and 99% of the events fall, respectively.

### Data Analysis and Figures

Frequency distribution histograms were created with 1-second intervals for apneas, hypopneas and cortical arousals, after which all arousals <3 or >30 seconds and all apneas or hypopneas <10 seconds or >90 seconds were removed. The 90-second cutoff value was selected because >99% apneas and hypopneas were shorter than this. The percentage of PSG records in each AHI severity category (<5, 5-15, 15-30, >30) and the mean ArI were calculated separately for each dataset.

Next, frequency distribution histograms for cortical arousal events vs. SDB events (apneas and hypopneas combined), apneas, and hypopneas were created by plotting the start time or the midpoint of an arousal event with respect to the end time of an SDB event within each recording. The distance between the start or the mid-point of an arousal event and the end of an SDB event was calculated for all possible arousal-SDB event pairs within a 60-second interval (from 30 seconds before until 30 seconds after the end of the SDB event) within each recording, and only the arousal event that was closest to a given SDB event end time was included. The two time points with a minimum number of arousal events on either side of the peak of the arousal distribution were identified and the cumulative distribution function (CDF) for the number of arousal events were calculated between these two local minimums across all data and separately for each dataset.

To explore differences in SDB-associated cortical arousal distributions between the datasets, a smoothed mean for a cortical arousal distribution (expressed as percentage of cortical arousal start times of the total number of cortical arousals within the selected 60-second interval) was calculated for each individual dataset, and the distributions were further divided into clinical research (647 records, 7 datasets) and community cohort (10,753 records, 3 datasets) distributions. A smoothed overall mean distribution was then calculated across the ten datasets (with each dataset contributing equally to the mean), along with a 95% confidence interval. To estimate how close together the arousal distributions cluster, we calculated the percentage of arousals for each individual distribution that fell between the two local minimums for the smoothed overall mean distribution.

To test the relationship between cortical arousals and SDB event duration, cortical arousal probability was calculated for different SDB event durations in 1-second resolution and plotted as a heatmap. Briefly, all SDB events were extracted and aligned such that the end of the event corresponded to time zero. All data from 10 seconds before until 90 seconds after the end of the SDB event were included. This interval was selected based on visual inspection of the data showing that very few arousal events start earlier than 10 seconds before or later than 20-30 seconds after the end of the SDB event, and that <1% of arousals were longer than 30 seconds. Each second of data within this interval was then classified into one of two dichotomous categories: arousal present or arousal absent (0), regardless of the arousal event start time or duration. Next, the SDB events were grouped into one-second bins based on event duration (i.e., first bin included all SDB events that were 10-seconds long, second bin included all SDB events that were 11-seconds long etc.). Arousal probability was calculated for each second within each bin by summing the total number of arousals present at a given second and dividing the number by the total number of SDB events in that bin. Finally, the resulting second-by-second cortical arousal probability for different SDB event durations was plotted as a heatmap, with arousal probability varying between 0% (arousal never present) to 100% (arousal always present). Similar heatmaps were generated for the clinical research and community cohort data and for each AHI severity category (<5, 5-15, 15-30, >30).

### Statistical Analysis

All statistical analyses were carried out in SAS (version 9.4). The relationship between AHI and ArI was explored using two linear mixed models; the first model used original data while the second model used standardized (log scaled) data. Both models were adjusted for apnea, hypopnea, and arousal mean event durations while considering the variability between the datasets (random effects). All outcomes were approximately normally distributed except AHI, which was log transformed for the second linear mixed model along with the rest of the data. The mean value (i.e., absolute number of arousals or arousal percentage within a selected time interval) was calculated separately for each dataset and across the data for each three outcomes (i.e., SDB events, apneas, and hypopneas). A smoothed curve was fitted individually to each dataset as well as to the overall mean across datasets using the penalized b-spline, and the 95% confidence intervals were calculated for the smoothed overall mean. Peak arousal probability was compared between nine different respiratory event duration categories (10<20 seconds, 20<30-seconds, 30<40 seconds, 40<50 seconds, 50<60 seconds, 60<70 seconds, 70<80 seconds, and 80<90 seconds) using a linear mixed model while considering the variability between the datasets (random effects). As there was very little variation in the location of the peak of the distribution between the different SDB event durations, we elected to compare the peak values rather than the area under curves or the quantiles. A p-value of <0.001 was considered statistically significant.

## RESULTS

We analyzed SDB events and cortical arousals from 11,400 PSG records that had been manually scored for SDB events and cortical arousals, originating from ten independent studies: from seven clinical sleep apnea studies and from three community cohort studies (Table 1). Together, these records included a total of 353,622 apneas (including central, mixed, and obstructive apneas), 1,979,606 hypopneas (including central, mixed, and obstructive hypopneas), and 1,626,480 cortical arousals (Supplemental Table S1).

To characterize the data, we calculated the AHI and the ArI for each record and the percentage of records in each AHI severity category and the mean ArI for each dataset (Table 1). We also correlated the AHI with the ArI across the 11,400 records and found that every 1.40-point increase (or every 1.54-point increase when using the log transformed data) in the AHI was associated with doubling of the ArI (original data: 1.40 with a 95% CI of [1.52, 1.57]; standardized data: 1.54 with a 95% CI of [1.39, 1.42]). Accoding to the frequency distribution histograms of the events data, 95% of all apneas, hypopneas, and cortical arousals were ≤47 seconds, ≤40 seconds and ≤23 seconds, respectively, while 99% of all apneas, hypopneas, and cortical arousals were ≤67 seconds, ≤61 seconds, and ≤30 seconds, respectively (Figure 1). Based on these data, we decided to include only those apneas (352,531) and hypopneas (1,974,880) that were ≥10 seconds and ≤90 seconds long and those cortical arousals (1,608,793) that were ≥3 seconds and ≤30 seconds long in further analysis. The respiratory events were analyzed both separately as apneas and hypopneas and as a group called SDB events (apneas and hypopneas combined).

Cortical arousals near the end of the respiratory events formed a roughly bell-shaped right skewed distribution with a distinct peak and two local minimums on either side of the peak. We considered a cortical arousal to be associated with an event if the start time of the arousal event fell within these two local minimums, i.e., between seconds -13 and +17 for SDB events (i.e., for all apneas and hypopneas combined, Figure 2 and Supplemental Table S2, upper panel) and for hypopneas (Figure 2 and Supplemental Table S2, lower panel), and between seconds -13 and +15 for apneas (Figure 2 and Supplemental Table S2, middle panel), as calculated across the entire dataset. Using this definition, we found that 90% of all SDB-associated cortical arousals started no earlier than 6 seconds before (5^th^ quantile) and no later than 14 seconds after (95^th^ quantile) the end of SDB event. These 5^th^ and 95^th^ quantile values were -4 seconds and +9 seconds for apnea-associated arousals, and -6 seconds and +14 seconds for hypopnea-associated arousals. The peak of the distribution coincided with the end of the respiratory event, i.e., the highest number of cortical arousals started at second 0 relative to the end of the event, regardless of event type. Similarly, we found that 90% of arousal midpoints occurred no earlier than 1 second before (5^th^ quantile) and no later than 20 seconds after (95^th^ quantile) the end of SDB events. These 5^th^ and 95^th^ quantile values were -1 second and +18 seconds for the combined apnea and hypopnea associated arousals (Figure 3, upper panel), +1 second and +15 seconds for apnea-associated arousals (Figure 3, middle panel), and -1 second and +20 seconds for hypopnea-associated arousals (Figure 3, lower panel). The peak of the distribution occurred approximately 6 seconds after the end of respiratory events, regardless of event type.

**Figure 2.**
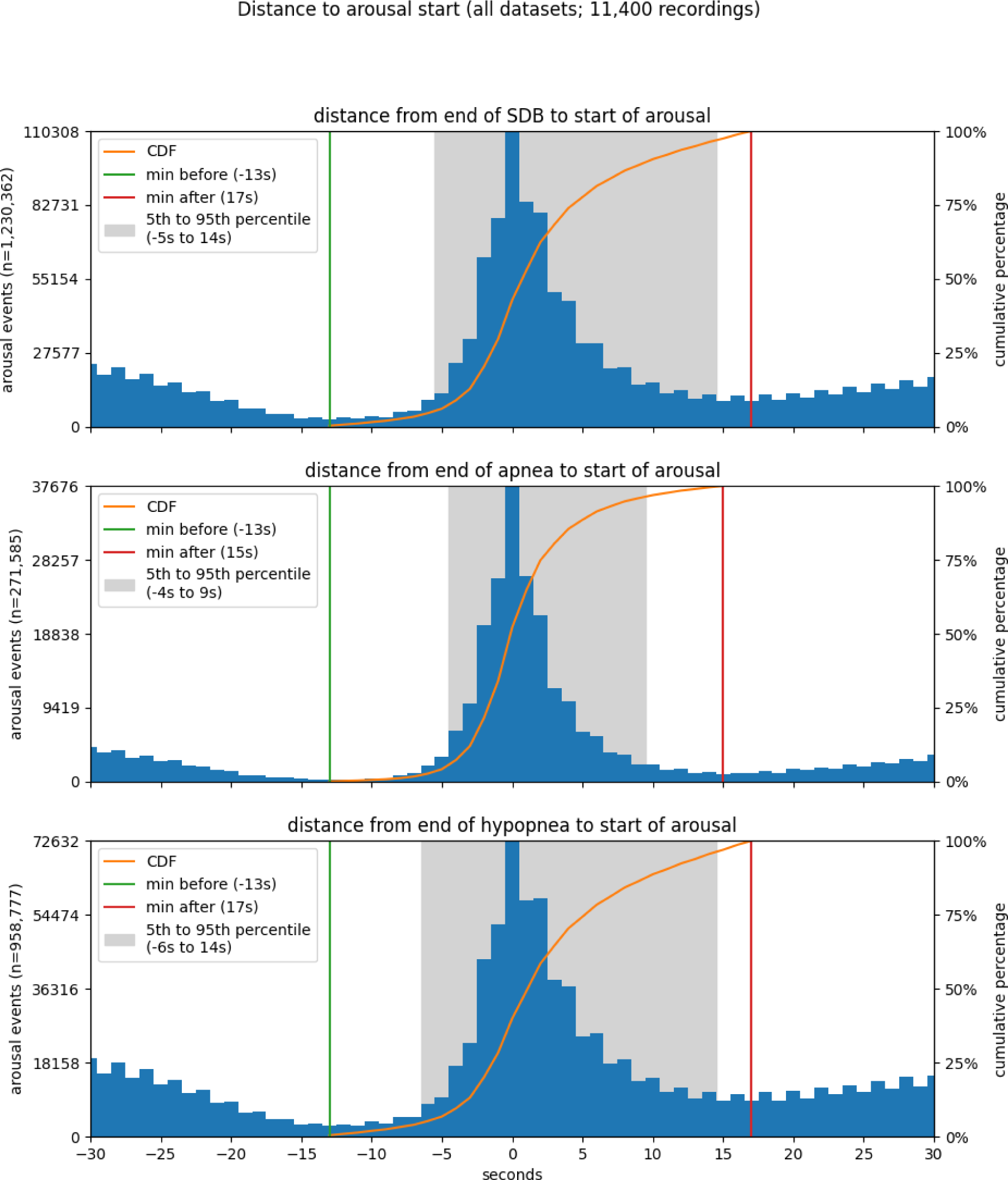
Frequency distribution histograms showing the total number per second (blue color, left y-axis) and the cumulative percentage between two local minimums (right y-axis) of cortical arousal start times relative to the end of SDB events (apneas and hypopneas combined, upper panel), apneas (middle panel), and hypopneas (lower panel) across all data (11,400 records). The x-axis ranges from 30 seconds before to 30 seconds after the end of the respiratory event, with 0 seconds corresponding to the end of the event. The two vertical lines indicate the time points with the fewest number of arousals starting before (green line) and after (red line) the peak of the distribution, the orange line shows the cumulative distribution function (CDF) for the percentage of arousals starting between the two lines, and the grey area indicates the area within which 90% (from 5^th^ to 95^th^ quantile) of these arousal start times fall.

**Figure 3.**
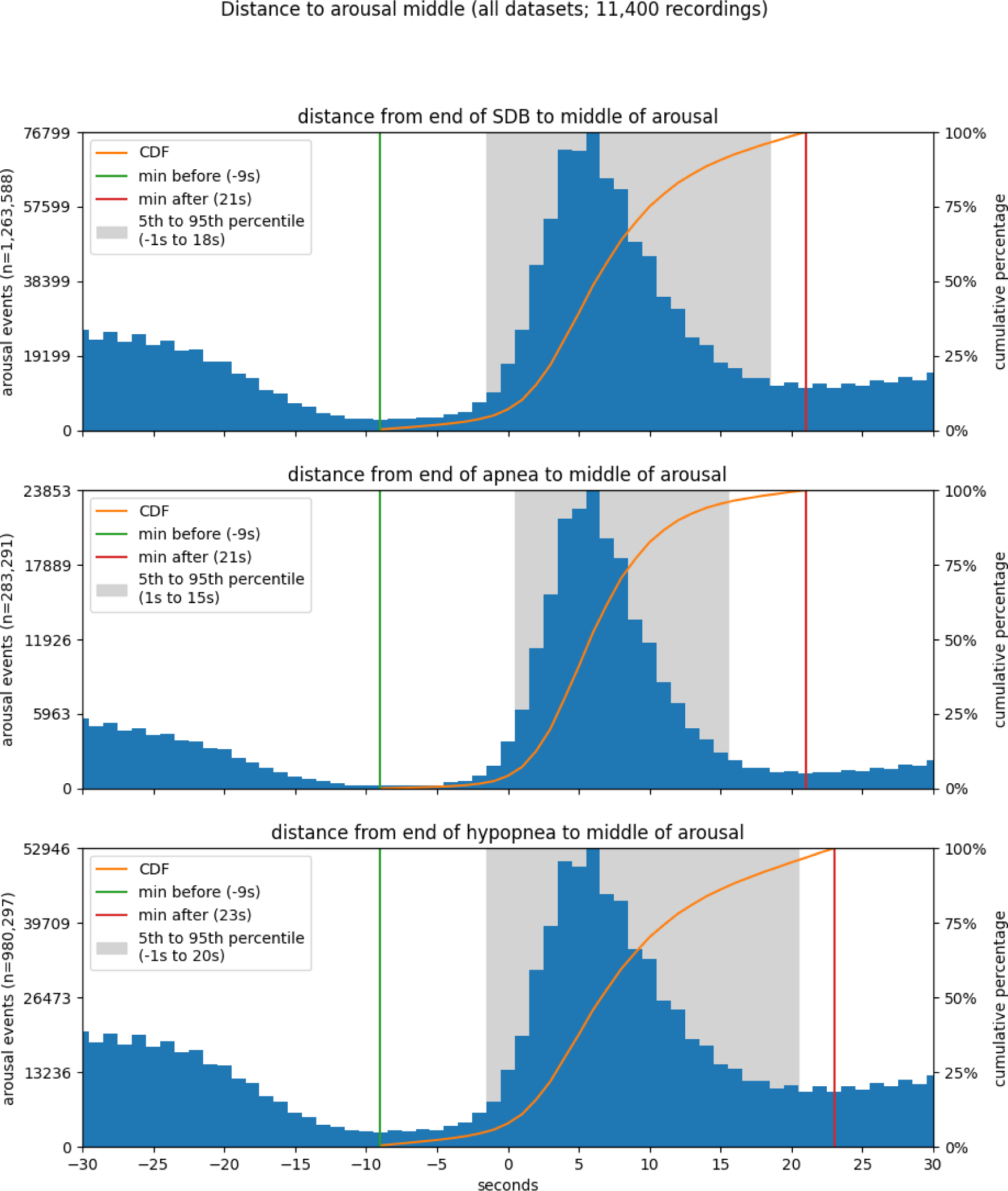
Frequency distribution histograms showing the total number per second (blue color, left y-axis) and the cumulative percentage between two local minimums (right y-axis) of cortical arousal midpoints relative to the end of SDB events (apneas and hypopneas combined, upper panel), apneas (middle panel), and hypopneas (lower panel) across all data (11,400 records). The x-axis ranges from 30 seconds before to 30 seconds after the end of the respiratory event, with 0 seconds corresponding to the end of the event. The two vertical lines indicate the time points with the fewest number of arousal midpoints before (green line) and after (red line) the peak of the distribution, the orange line shows the cumulative distribution function (CDF) for the percentage of arousal midpoints between the two lines, and the grey area indicates the area within which 90% (from 5^th^ to 95^th^ quantile) of the arousal midpoints fall.

To explore potential differences in the timing of SDB-associated cortical arousals between the datasets, we compared the individual arousal distributions (Supplemental Table S3) to the smoothed overall mean distribution calculated across the ten datasets. The peak of the smoothed mean cortical arousal distribution occurred one second before the end of the events, and the two local minimums occurred 13 seconds before and 12 seconds after the end of the events, regardless of event type (SDB events, Figure 4, upper panel; apneas, Figure 4, middle panel; hypopneas, Figure 4, lower panel). This was similar to the unsmoothed data (Figure 2) where the peak of the distribution occurred at second 0 before the end of the respiratory events and the two local minimums occurred 13 seconds before and 17 seconds after (15 seconds after for apneas) the end of the respiratory events. These local minimums as well as the 1^th^-10^th^ and 90^th^-99^th^ quantile values for the unsmoothed SDB-associated arousal distributions for each dataset are reported in Supplemental Table S2. The percentage of cortical arousals (within the selected 60-second interval) that started at the peak of the smoothed mean distribution was 11.0% (95% CI [7.6-14.3]) for arousals following SDB events, 12.9% [9.8-16.0] for arousals following apneas, and 10.1% [6.8-13.5] for arousals following hypopneas (Figure 4). While there was variability in the shape of the individual distributions, they clustered around the overall mean distribution and were either within, or close, to the 95% confidence interval for the overall mean (Figure 4). However, the distributions for the community cohort studies (MESA, SHHS, SOF) were shifted to right compared to the overall mean distribution, whereas the distributions for the clinical research studies (ABC, ATS, MPCS, CTS, CHFS, MSS, TS) were close to the overall mean distribution or slightly shifted to the left (Figure 4). Nevertheless, for both community and clinical studies, the distance from the end of an SDB event to the start of an arousal fell between the two local minimums of the smoothed overall mean distribution across all 10 data sets for ≥90% of the apnea and hypopneas events.

**Figure 4.**
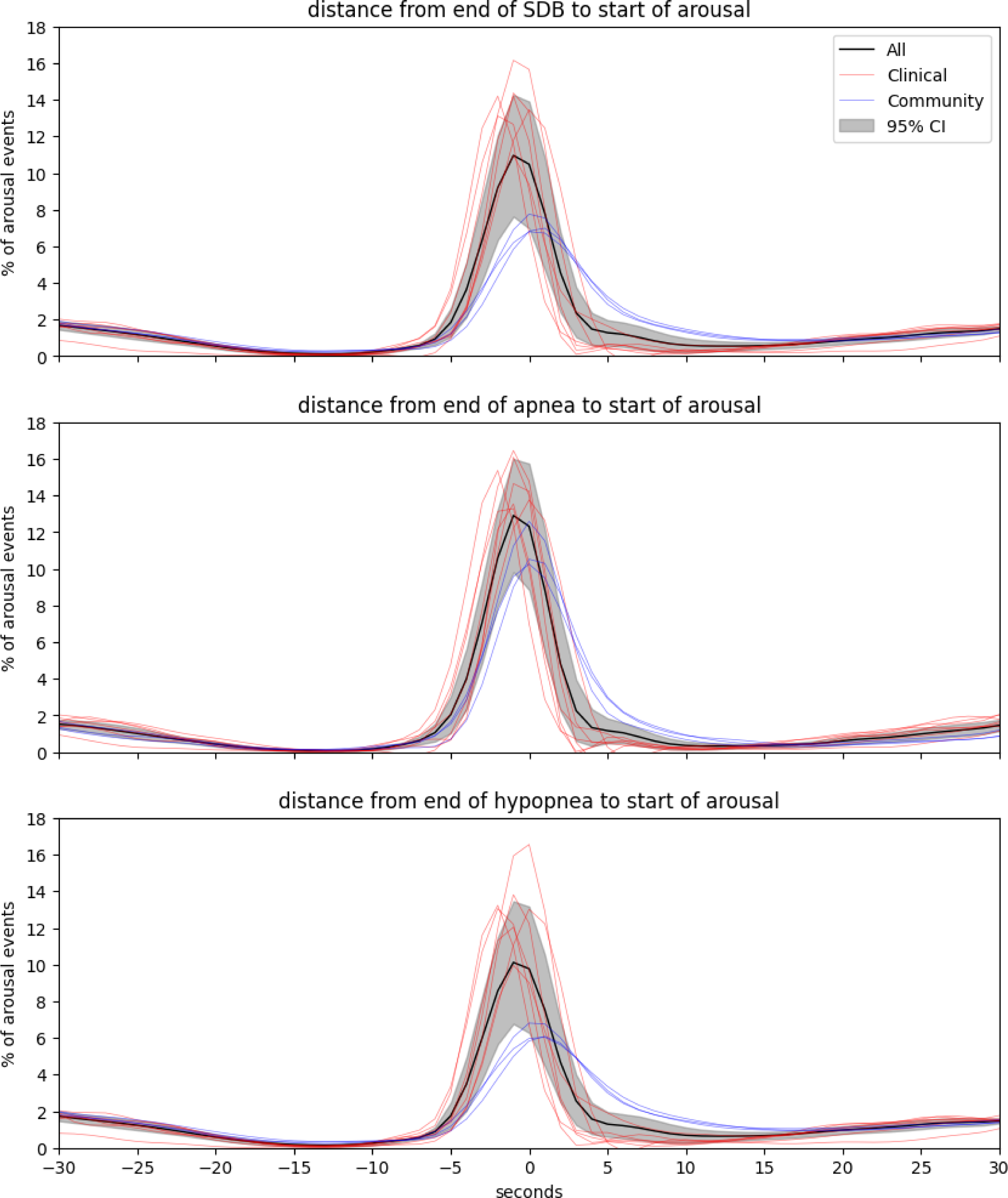
Frequency distribution histograms showing the number cortical arousal start times relative to the end of SDB events (apneas and hypopneas combined, upper panels), apneas (middle panels), and hypopneas (lower panels) across the data. Black line shows the smoothed overall mean across the ten datasets, while the grey color indicates the upper and lower bounds for the 95% confidence interval. The x-axis ranges from 30 seconds before to 30 seconds after the end of the respiratory event, with 0 seconds corresponding to the end of the event. The red lines show smoothed data for the seven clinical research datasets and the blue lines show soothed data for the three community cohort datasets (see Supplemental Table S3 for data for the individual datasets). The y-axis shows cortical arousal start times expressed as percentage of the total number of cortical arousal start times within the selected 60-second interval (the Area Under Curve for each line is 100%).

Cortical arousal probability (i.e., the likelihood for a cortical arousal to be present at a given second) was highest during the first 10 seconds after the end of an SDB event (Figure 5, upper panel heatmap), apnea (Figure 5, middle panel heatmap), or hypopnea (Figure 5, lower panel heatmap). While there was no major shift in the location of peak of the distribution, which occurred approximately 5 seconds after the end of the respiratory event regardless of event type or duration, there was a significant difference in arousal probability between different SDB event durations (Figure 5; p<0.0001 for SDB events, p<0.0001 for apneas, and p<0.0001 for hypopneas), such that arousals were more frequent following longer SDB events compared to shorter SDB events. The estimated (least square means) arousal peak probability was 43.6%, 54.0%, 62.2%, 65.1%, 65.5%, 63.5%, 63.7%, and 64.2% for SDB events that were 10<20-seconds, 20<30-seconds, 30<40-seconds, 40<50 seconds, 50<60 seconds, 60<70 seconds, 70<80 seconds, and 80<90 seconds long, respectively (Figure 5, upper panel). These estimates were 46.7%, 63.1%, 73.7%, 77.7%, 79.7%, 77.0%, 82.7% and 81.6% for apnea-associated arousals (Figure 5 middle panel), and 46.6%, 50.9%, 55.5%, 56.9%, 57.0%, 56.6%, 59.3% and 62.1% for hypopnea-associated arousals (Figure 5, lower panel). Finally, as expected, cortical arousal probability was higher in the clinical research datasets compared to the community cohort datasets (Supplemental Figure S1), and also higher with higher AHI (Supplemental Figure S2).

**Figure 5.**
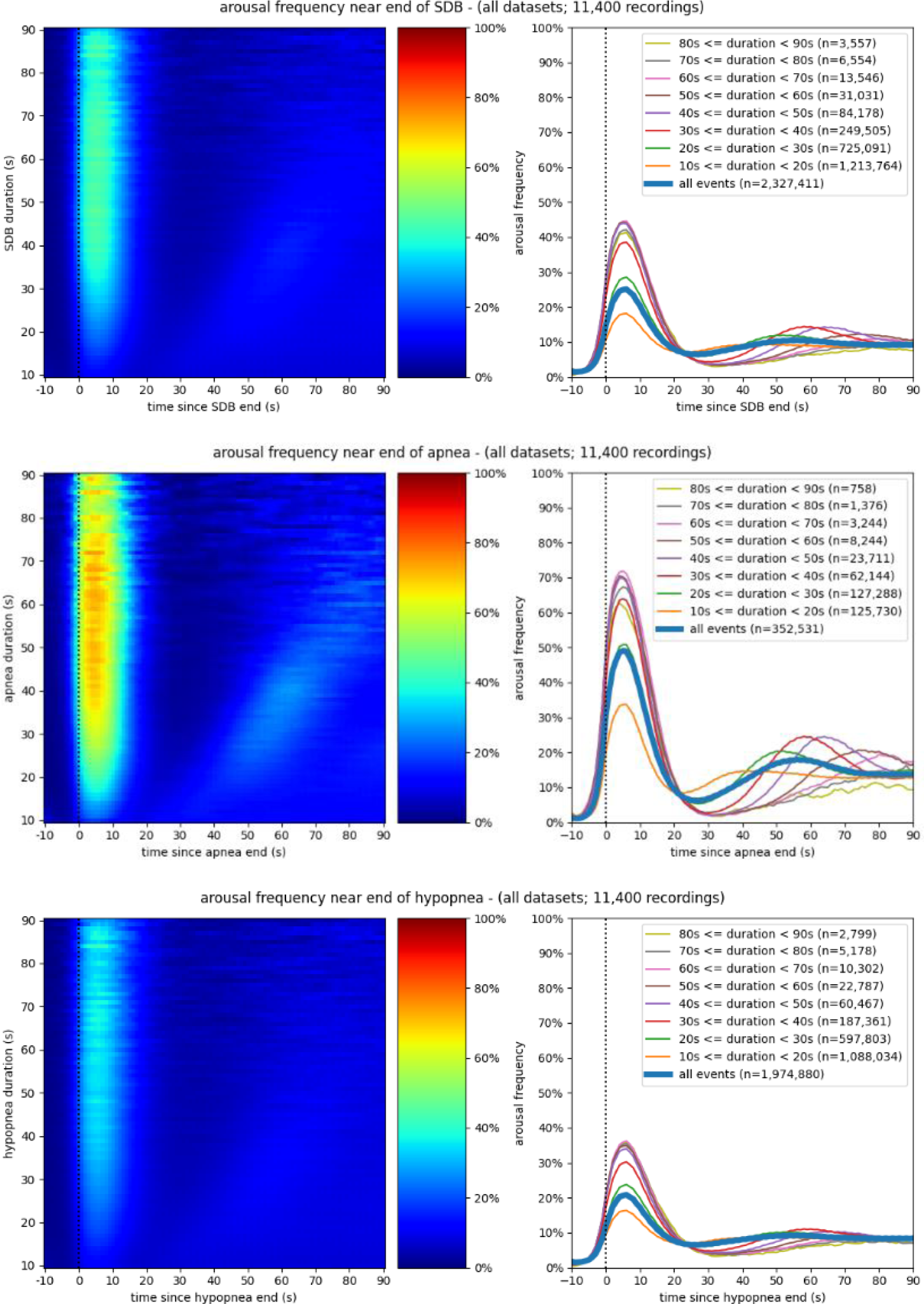
Heatmaps showing cortical arousal probability as a function of SDB (apneas and hypopneas combined, upper panel), apnea (middle panel), and hypopnea (lower panel) event duration and time since the end of the event. X-axis indicates time since the end of the event and ranges from 10 seconds before to 90 seconds after the end of the event, with 0 corresponding to the end of the event. Y-axis indicates event duration and ranges from 10 seconds to 90 seconds. Color on the heatmap indicates cortical arousal probability in 1-second resolution (i.e., the likelihood of an arousal to be present at a given second) and ranges from 0% (dark blue) to 100% (dark red) and the color scales have been standardized between the panels to make data comparable across event categories. Arousal probability curves next to the heatmaps are a quantification of the data in the heatmap and indicate arousal probability (y-axis) for each second of the data (x-axis) for a given SDB event duration (line color).

## DISCUSSION

The AASM recommends scoring hypopneas in adults when there is a ≥3% oxygen desaturation or when the event is associated with an arousal,^9^ but there is no rule regarding the duration of the interval between an event and the associated arousal. We explored the relationship between cortical arousals and SDB events in 11,400 adult PSG records from ten independent clinical research and community cohort datasets that had been manually scored for apneas, hypopneas, and cortical arousals.

There were more than 2.3 million scored SDB events (>350,000 apneas and >1.9 million hypopneas, including central, mixed, and obstructive events) and more than 1.6 million cortical arousals in the datasets. Approximately 99% of all SDB events were shorter than 60 seconds and 99% of all cortical arousals were shorter than 30 seconds. The Apnea, Bariatric Surgery, and CPAP (ABC) study^11^ had the highest number of records in the most severe AHI>30 category (54.5%), while the Study of Osteoporotic Fractures (SOF) study^11^ had the fewest (10.7%). Similarly, the Taranto Muscle Stimulation Study (MSS)^19^ had the highest number of records in the least severe AHI<5 category (36%), while the ABC study had the fewest (0.8%). As expected, AHI correlated positively with the ArI, such that the AHI increased by 1.40-1.54 points with each doubling of the ArI. This is in line with previous studies that have found strong correlation between AHI and ArI.^22^

Cortical arousals that started near the end of the respiratory events formed a bell-shaped right skewed distribution, with a distinct peak and two local minimums on either side of the peak. A cortical arousal was considered to be associated with an event if the start time of the arousal fell within these two local minimums. Using this definition, we found that 90% of all SDB (i.e., apnea and hypopnea)-associated cortical arousals started within a 20-second interval that started no earlier than 6 seconds before and no later than 14 seconds after the end of the SDB event, with the peak of the distribution coinciding with the end of the respiratory event. Cortical arousals associated with apneas were more tightly clustered around the end of the apnea event (within a 13-second interval) compared to cortical arousals associated with hypopneas (within a 20-second interval), which may be due to apneas being associated with greater oxygen desaturation and greater physiologic stress compared to hypopneas. Similarly, we found that 90% of cortical arousal midpoints associated with SDB (apnea and hypopnea) events occurred within a 21-second interval that started no earlier than 1 second before and no later than 20 seconds after the end of the SDB event, with the peak of the distribution occurring at 6 seconds after the end of the event.

We also compared cortical arousal distributions from different datasets to the smoothed overall mean distribution. In clinical studies, the scorer typically views all the data simultaneously and uses the presence of an arousal/desaturation as a clue to the occurrence of the event, while in community studies, a change in flow or effort is often identified, and a software algorithm may be used to match the presence of an arousal/desaturation to the event. Hypopnea events in two of the community cohort studies, the SHHS^21^ and the MESA,^23^ were scored using a decrease in respiratory signal or effort rather than the presence of an arousal/desaturation. Thus, these two datasets, which together account for 92% of the SDB events, present a less biased view on the association between arousals and SDB events compared to the other datasets, which used arousals/desaturations for hypopnea scoring. However, while the arousal distributions were slightly different between the community and clinical studies, with the distributions for the community datasets wider and shifted to right while the distributions for the clinical datasets were narrower and closer to the mean or shifted to the left, there was a substantial overlap between the distributions. Moreover, similar differences between the clinical and community datasets were observed for arousal distributions associated with apneas (which, unlike hypopneas, are not scored using the presence of an arousal). Finally, the third community cohort dataset, the SOF,^11^ whose SDB-associated arousal distributions were almost identical to those of the SHHS and MESA, did use the presence of arousals in SDB scoring. Together, these results suggest that differences in the timing of SDB-associated arousals between the datasets are small and may be due to differences in data collection rather than differences in event scoring.

The peak of the smoothed overall mean distribution for cortical arousals (calculated across datasets) occurred one second before the end of the events, and the local minimums occurred 13 seconds before and 12 seconds after the end of the respiratory events, regardless of event type. This is similar to the unsmoothed mean distribution for cortical arousals where the peak of the SDB-associated arousal distribution occurred at second 0 relative to the end of the respiratory events, and the local minimums occurred 13 seconds before and 17 seconds (15 seconds for apnea events) after the end of the SDB events.

Finally, we found that cortical arousal probability near the end of respiratory events was highest during the first 10 seconds after the end of the event, with a peak around 5 seconds after the end of the event, regardless of event type or duration. This is likely because most cortical arousals are short in duration and because, as described above, SDB event-associated cortical arousals tend to start very close to the end of the respiratory events. However, cortical arousal probability near the end of SDB events was not uniform but varied by event type and duration and by AHI severity. Cortical arousal probability was higher for longer respiratory events compared to shorter events, it was higher following apnea events compared to hypopnea events, and it increased with increasing AHI severity. These findings are in line with some, although not all, previous studies showing that while the average arousal intensity is not related to the magnitude of the preceding respiratory stimuli^24,25^, there is an increased cortical response to apneas compared to hypopneas.^26-29^ Future studies are needed to explore the role of arousals in breathing-related sleep disorders.

In summary, we found that cortical arousals near the end of SDB events form a bell-shaped right skewed distribution and that 90% of arousals within this distribution start no earlier than 6 seconds before (5^th^ quantile) and no later than 14 seconds after (95^th^ quantile) the end of the SDB event, with the peak of the arousal distribution coinciding with the end of the SDB event. These data should be considered when developing a more precise definition for the association between cortical arousals and SDB events.

## Data Availability

Some of the data are publicly available at the National Sleep Research Resource (NSRR). Execution of a materials transfer agreement is required by our institution for transfer of the non-publicly available data.

https://sleepdata.org/

