## Supplemental File for "Association of cortical arousals with sleep-disordered breathing events"

### Supplemental Material

| original label | new label | total | SHHS | MESA | ABC | SOF | ATS | MPCS | CTS | CHFS | MSS | TS |
|---|---|---|---|---|---|---|---|---|---|---|---|---|
| 2 | sleep | 4628490 | 3545240 | 854824 | 52334 | 176092 | | | | | | |
| 0 | wake | 4285388 | 2899614 | 1127878 | 30938 | 226958 | | | | | | |
| Hypopnea | hypopnea | 1901249 | 1588062 | 230202 | 21428 | 61557 | | | | | | |
| SpO2 desaturation | | 1885773 | 1151606 | 680622 | 53328 | 217 | | | | | | |
| 5 | sleep | 1574398 | 1228628 | 268799 | 18671 | 58300 | | | | | | |
| Arousal () | arousal | 1363508 | 1072554 | 257267 | 31790 | 1897 | | | | | | |
| 3 | sleep | 1214700 | 993260 | 149464 | 11761 | 60215 | | | | | | |
| 1 | sleep | 568170 | 329039 | 203957 | 19296 | 15878 | | | | | | |
| SpO2 artifact | | 331089 | 286898 | 24948 | 2042 | 17201 | | | | | | |
| Obstructive Apnea | apnea | 302162 | 238656 | 46686 | 7539 | 9281 | | | | | | |
| S2 | sleep | 205637 | | | | | 51246 | 19552 | 36590 | 19635 | 67083 | 11531 |
| W | wake | 187613 | | | | | 46576 | 17196 | 18714 | 27418 | 67389 | 10320 |
| PLM (Left) | | 182347 | | 170618 | 11479 | 250 | | | | | | |
| Unsure | hypopnea | 177838 | 1219 | 164961 | 11658 | | | | | | | |
| AR | arousal | 142466 | | | | | 25404 | 15803 | 13764 | 12071 | 64608 | 10816 |
| Limb Movement (Left) | | 126414 | | 59713 | 1760 | 64941 | | | | | | |
| Arousal (ASDA) | arousal | 123407 | 3429 | 61955 | | 58023 | | | | | | |
| S1 | sleep | 90356 | | | | | 17515 | 7731 | 7388 | 10175 | 42075 | 5472 |
| Limb Movement (Right) | | 72080 | | | 2800 | 69280 | | | | | | |
| 4 | sleep | 64369 | 59933 | 210 | | 4226 | | | | | | |
| REM | sleep | 46182 | | | | | 14291 | 5254 | 8204 | 3293 | 12410 | 2730 |
| Central Apnea | apnea | 45771 | 38217 | 5825 | 651 | 1078 | | | | | | |
| HypO | hypopnea | 36727 | | | | | 5360 | 3346 | 5642 | 1843 | 15525 | 5011 |
| ApO | apnea | 25841 | | | | | 5759 | 2690 | 4409 | 2486 | 7175 | 3322 |
| S3 | sleep | 24748 | | | | | 4038 | 4059 | 1828 | 365 | 11636 | 2822 |
| PLM (Right) | | 15775 | | | 15422 | 353 | | | | | | |
| Arousal (STANDARD) | arousal | 12345 | 12345 | | | | | | | | | |
| H | hypopnea | 8474 | | | | | 3382 | | 2716 | 2376 | | |
| ? | | 7154 | | | | | 2103 | 458 | 72 | 661 | 3860 | |
| ap-O | apnea | 6768 | | | | | 4053 | | 1457 | 1258 | | |
| no stage | | 4169 | | | | | 2999 | 22 | 86 | 970 | 67 | 25 |
| M | apnea | 4068 | | | | | 1459 | 96 | 645 | 1018 | 381 | 469 |
| ApC | apnea | 3533 | | | | | 1113 | 485 | 399 | 710 | 632 | 194 |
| Arousal (ARO SPONT) | arousal | 3110 | | 3110 | | | | | | | | |
| HypOx | hypopnea | 1213 | | | | | | | 944 | 266 | 2 | 1 |
| U | | 1153 | | | | | 965 | 36 | 1 | 1 | 137 | 13 |
| Arousal (Standard) | arousal | 1103 | 1103 | | | | | | | | | |
| Ap-C | apnea | 1041 | | | | | 856 | | 71 | 114 | | |
| Arousal (CHESHIRE) | arousal | 980 | 980 | | | | | | | | | |
| HypC | hypopnea | 945 | | | | | 133 | | 2 | 809 | 1 | |
| Arousal (ARO Limb) | arousal | 938 | | 938 | | | | | | | | |
| Mixed Apnea | apnea | 808 | 530 | 276 | | 2 | | | | | | |
| Arousal (asda) | arousal | 675 | | | | 675 | | | | | | |
| 9 | | 471 | 254 | 89 | 101 | 27 | | | | | | |
| Arousal (ARO RES) | arousal | 453 | 111 | 145 | 197 | | | | | | | |
| HypO2% | hypopnea | 374 | | | | | | | 232 | 141 | | 1 |
| Hyp-C | hypopnea | 309 | | | | | | | | 309 | | |
| Arousal (AASM) | arousal | 177 | | 177 | | | | | | | | |
| HypCx | hypopnea | 174 | | | | | | | | 174 | | |
| Arousal (Arousal) | arousal | 157 | | | | 157 | | | | | | |
| Arousal (External Arousal) | arousal | 120 | 120 | | | | | | | | | |
| HypC2% | hypopnea | 61 | | | | | | | | 61 | | |
| Arousal (RESP ARO) | arousal | 26 | | | | 26 | | | | | | |
| Respiratory artifact | | 25 | 7 | 15 | 1 | 2 | | | | | | |
| RERA | arousal | 13 | | 13 | | | | | | | | |
| HypO ??? | | 6 | | | | | 6 | | | | | |
| 6 | | 3 | 3 | | | | | | | | | |
| Proximal pH artifact | | 2 | | | 2 | | | | | | | |
| Periodic Breathing | | 2 | 1 | 1 | | | | | | | | |
| TcCO2 artifact | | 1 | | | 1 | | | | | | | |
| Blood pressure artifact | | 1 | | | 1 | | | | | | | |
| Arousal (PLM ARO) | arousal | 1 | | | | 1 | | | | | | |
| Arousal (Cheshire) | arousal | 1 | 1 | | | | | | | | | |
| Body temperature artifact | | 1 | | | 1 | | | | | | | |
| ApC ??? | | 1 | | | | | 1 | | | | | |
| ApO ??? | | 1 | | | | | 1 | | | | | |
| ApC ??? | | 1 | | | | | 1 | | | | | |
| Distal pH | | 1 | | | 1 | | | | | | | |
| HypO ??? | | 1 | | | | | 1 | | | | | |
| Distal pH artifact | | 1 | | | 1 | | | | | | | |
| EtCO2 artifact | | 1 | | | 1 | | | | | | | |
| test | | 1 | | | | | | | | | 1 | |

Table S1. Events by dataset. Original label indicates how the event was originally labelled while the new label indicates how the event was relabeled for analysis. The total number of events in the table (19,683,361) includes all events and event types from the manually scored output files before relabeling the events, filtering out duplicate events, and applying overlap correction.

| | min before | quantiles (%) for SDB-associated cortical arousal distributions | | | | | | | | | | | | | | | | | | | | min after |
|---|---|---|---|---|---|---|---|---|---|---|---|---|---|---|---|---|---|---|---|---|---|---|
| | | 1% | 2% | 3% | 4% | 5% | 6% | 7% | 8% | 9% | 10% | 90% | 91% | 92% | 93% | 94% | 95% | 96% | 97% | 98% | 99% | |
| ALL | -13 | -11 | -9 | -7 | -6 | -5 | -5 | -4 | -4 | -3 | -3 | 10 | 11 | 12 | 12 | 13 | 14 | 14 | 15 | 16 | 17 | 17 |
| SHHS | -11 | -9 | -8 | -6 | -5 | -5 | -4 | -4 | -3 | -3 | -3 | 11 | 11 | 12 | 13 | 13 | 14 | 15 | 15 | 16 | 17 | 17 |
| MESA | -13 | -10 | -8 | -7 | -6 | -5 | -5 | -4 | -4 | -4 | -3 | 11 | 12 | 13 | 14 | 14 | 15 | 16 | 17 | 18 | 19 | 19 |
| SOF | -13 | -11 | -9 | -7 | -6 | -6 | -5 | -4 | -4 | -4 | -4 | 13 | 14 | 14 | 15 | 16 | 17 | 18 | 19 | 20 | 20 | 21 |
| ABC | -12 | -8 | -6 | -6 | -5 | -5 | -4 | -4 | -4 | -4 | -3 | 6 | 7 | 7 | 8 | 9 | 10 | 11 | 12 | 13 | 14 | 15 |
| ATS | -13 | -6 | -5 | -5 | -4 | -4 | -4 | -3 | -3 | -3 | -3 | 2 | 2 | 3 | 3 | 4 | 4 | 5 | 6 | 7 | 10 | 13 |
| MPCS | -12 | -9 | -7 | -7 | -6 | -5 | -5 | -5 | -4 | -4 | -4 | 1 | 1 | 2 | 2 | 2 | 3 | 4 | 5 | 6 | 7 | 9 |
| CST | -14 | -9 | -8 | -7 | -6 | -6 | -6 | -5 | -5 | -5 | -5 | 1 | 1 | 1 | 2 | 2 | 2 | 3 | 4 | 6 | 8 | 11 |
| CHFS | -19 | -8 | -7 | -6 | -5 | -5 | -5 | -4 | -4 | -4 | -4 | 2 | 3 | 3 | 4 | 4 | 5 | 6 | 8 | 11 | 14 | 18 |
| MSS | -12 | -8 | -7 | -6 | -5 | -5 | -4 | -4 | -4 | -4 | -3 | 1 | 2 | 2 | 2 | 3 | 3 | 4 | 5 | 6 | 7 | 9 |
| TS | -14 | -10 | -8 | -8 | -7 | -6 | -6 | -6 | -5 | -5 | -5 | 0 | 0 | 1 | 1 | 1 | 1 | 2 | 3 | 4 | 6 | 10 |

| | min before | quantiles (%) for apnea-associated cortical arousal distributions | | | | | | | | | | | | | | | | | | | | min after |
|---|---|---|---|---|---|---|---|---|---|---|---|---|---|---|---|---|---|---|---|---|---|---|
| | | 1% | 2% | 3% | 4% | 5% | 6% | 7% | 8% | 9% | 10% | 90% | 91% | 92% | 93% | 94% | 95% | 96% | 97% | 98% | 99% | |
| ALL | -13 | -8 | -6 | -5 | -5 | -4 | -4 | -4 | -3 | -3 | -3 | 6 | 6 | 7 | 7 | 8 | 9 | 10 | 11 | 12 | 14 | 15 |
| SHHS | -13 | -8 | -6 | -5 | -4 | -4 | -3 | -3 | -3 | -3 | -2 | 7 | 7 | 8 | 8 | 9 | 9 | 10 | 11 | 12 | 14 | 15 |
| MESA | -17 | -11 | -8 | -6 | -6 | -5 | -5 | -4 | -4 | -4 | -3 | 6 | 6 | 6 | 7 | 8 | 8 | 9 | 11 | 12 | 15 | 17 |
| SOF | -15 | -10 | -8 | -7 | -6 | -5 | -5 | -4 | -4 | -4 | -3 | 8 | 9 | 9 | 10 | 11 | 12 | 14 | 15 | 17 | 19 | 21 |
| ABC | -15 | -8 | -7 | -6 | -6 | -6 | -5 | -5 | -5 | -4 | -4 | 2 | 2 | 3 | 3 | 4 | 4 | 5 | 5 | 7 | 8 | 11 |
| ATS | -14 | -6 | -5 | -4 | -4 | -4 | -3 | -3 | -3 | -3 | -3 | 2 | 2 | 2 | 3 | 3 | 4 | 4 | 5 | 7 | 9 | 13 |
| MPCS | -13 | -8 | -7 | -6 | -5 | -5 | -4 | -4 | -4 | -3 | -3 | 1 | 1 | 1 | 2 | 2 | 3 | 4 | 5 | 6 | 7 | 9 |
| CST | -14 | -9 | -8 | -7 | -7 | -6 | -6 | -6 | -5 | -5 | -5 | 0 | 1 | 1 | 1 | 1 | 2 | 3 | 3 | 5 | 7 | 10 |
| CHFS | -19 | -8 | -6 | -6 | -5 | -5 | -5 | -4 | -4 | -4 | -4 | 2 | 2 | 2 | 2 | 3 | 3 | 4 | 5 | 6 | 8 | 11 |
| MSS | -12 | -7 | -6 | -5 | -5 | -4 | -4 | -4 | -3 | -3 | -3 | 1 | 1 | 1 | 2 | 2 | 2 | 3 | 4 | 6 | 8 | 11 |
| TS | -12 | -9 | -8 | -7 | -7 | -6 | 6 | -5 | -5 | -5 | -5 | 0 | 0 | 1 | 1 | 1 | 2 | 2 | 4 | 5 | 6 | 9 |

| | min before | quantiles (%) for hypopnea-associated cortical arousal distributions | | | | | | | | | | | | | | | | | | | | min after |
|---|---|---|---|---|---|---|---|---|---|---|---|---|---|---|---|---|---|---|---|---|---|---|
| | | 1% | 2% | 3% | 4% | 5% | 6% | 7% | 8% | 9% | 10% | 90% | 91% | 92% | 93% | 94% | 95% | 96% | 97% | 98% | 99% | |
| ALL | -13 | -11 | -9 | -8 | -6 | -6 | -5 | -4 | -4 | -4 | -3 | 11 | 12 | 12 | 13 | 14 | 14 | 15 | 16 | 16 | 17 | 17 |
| SHHS | -11 | -10 | -8 | -7 | -6 | -5 | -4 | -4 | -4 | -3 | -3 | 12 | 12 | 13 | 13 | 14 | 14 | 15 | 16 | 16 | 17 | 17 |
| MESA | -13 | -11 | -9 | -7 | -6 | -5 | -5 | -4 | -4 | -4 | -3 | 12 | 13 | 14 | 14 | 15 | 16 | 17 | 17 | 18 | 19 | 19 |
| SOF | -13 | -11 | -9 | -8 | -7 | -6 | -5 | -5 | -4 | -4 | -4 | 11 | 12 | 12 | 13 | 13 | 14 | 15 | 15 | 16 | 17 | 17 |
| ABC | -12 | -8 | -6 | -5 | -5 | -4 | -4 | -4 | -3 | -3 | -3 | 6 | 7 | 7 | 8 | 8 | 9 | 10 | 11 | 12 | 12 | 13 |
| ATS | -13 | -7 | -6 | -5 | -5 | -4 | -4 | -4 | -3 | -3 | -3 | 2 | 2 | 3 | 3 | 4 | 4 | 5 | 6 | 7 | 8 | 10 |
| MPCS | -12 | -9 | -8 | -7 | -6 | -6 | -5 | -5 | -5 | -4 | -4 | 1 | 2 | 2 | 2 | 2 | 3 | 4 | 5 | 6 | 8 | 10 |
| CST | -14 | -10 | -8 | -7 | -6 | -6 | -6 | -5 | -5 | -5 | -5 | 1 | 1 | 1 | 2 | 2 | 3 | 3 | 5 | 6 | 8 | 11 |
| CHFS | -19 | -8 | -7 | -6 | -5 | -5 | -4 | -4 | -4 | -4 | -3 | 3 | 3 | 4 | 4 | 5 | 5 | 6 | 8 | 10 | 12 | 15 |
| MSS | -12 | -8 | -7 | -6 | -6 | -5 | -5 | -4 | -4 | -4 | -4 | 2 | 2 | 2 | 3 | 3 | 4 | 4 | 5 | 6 | 8 | 9 |
| TS | -14 | -10 | -8 | -7 | -7 | -6 | -6 | -6 | -5 | -5 | -5 | 0 | 0 | 1 | 1 | 1 | 1 | 1 | 2 | 3 | 4 | 7 |

Table S2. The two local minimums (min before, min after) and the $1^{th}$-$10^{th}$ and $90^{th}$-$99^{th}$ quantiles (%) for the unsmoothed SDB, apnea, and hypopnea-associated cortical arousal distribution across all data (ALL) and for each individual dataset (SHHS, MESA, SOF, ABC, ATS, MPCS, CST, CHFS, MSS, TS). The values are in seconds and range from a minimum value of -19 (i.e., 19 seconds before the end of the respiratory event) to a maximum value of 21 (i.e., 21 seconds after the end of the respiratory event).

Table S3. Smoothed mean cortical arousal start time distributions (expressed as percentage of the total number of cortical arousal start times within the selected 60-second interval) for each of the ten datasets.

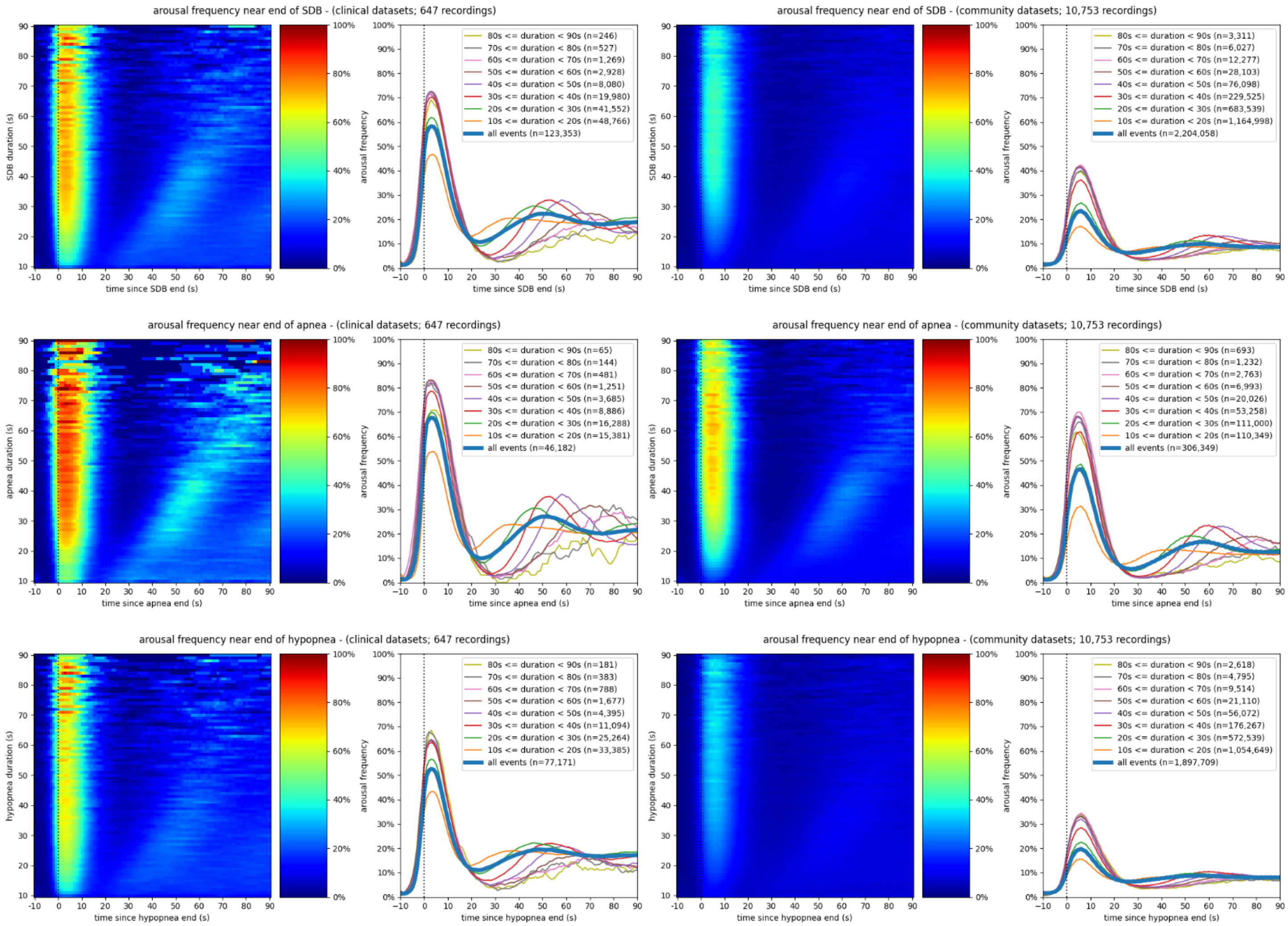


Figure S1. Heatmap showing cortical arousal probability as a function of SDB (apneas and hypopneas combined, upper panel), apnea (middle panel), and hypopnea (lower panel) event duration and time since the end of the event for clinical datasets (left side panels) and for community datasets (right side panels). X-axis indicates time since the end of the event and ranges from 10 seconds before to 90 seconds after the end of the event, with 0 corresponding to the end of the event. Y-axis shows event duration and ranges from 10 seconds to 90 seconds. Color on the heatmap indicates cortical arousal probability in 1-second resolution (i.e., the likelihood of an arousal to be present at a given second) and ranges from 0% (dark blue) to 100% (dark red) and the color scales have been standardized between the panels to make data comparable across event categories. Arousal probability curves next to the heatmaps are a quantification of the data in the heatmaps and indicate arousal probability (y-axis) for each second of the data (x-axis) for a given event duration (line color).

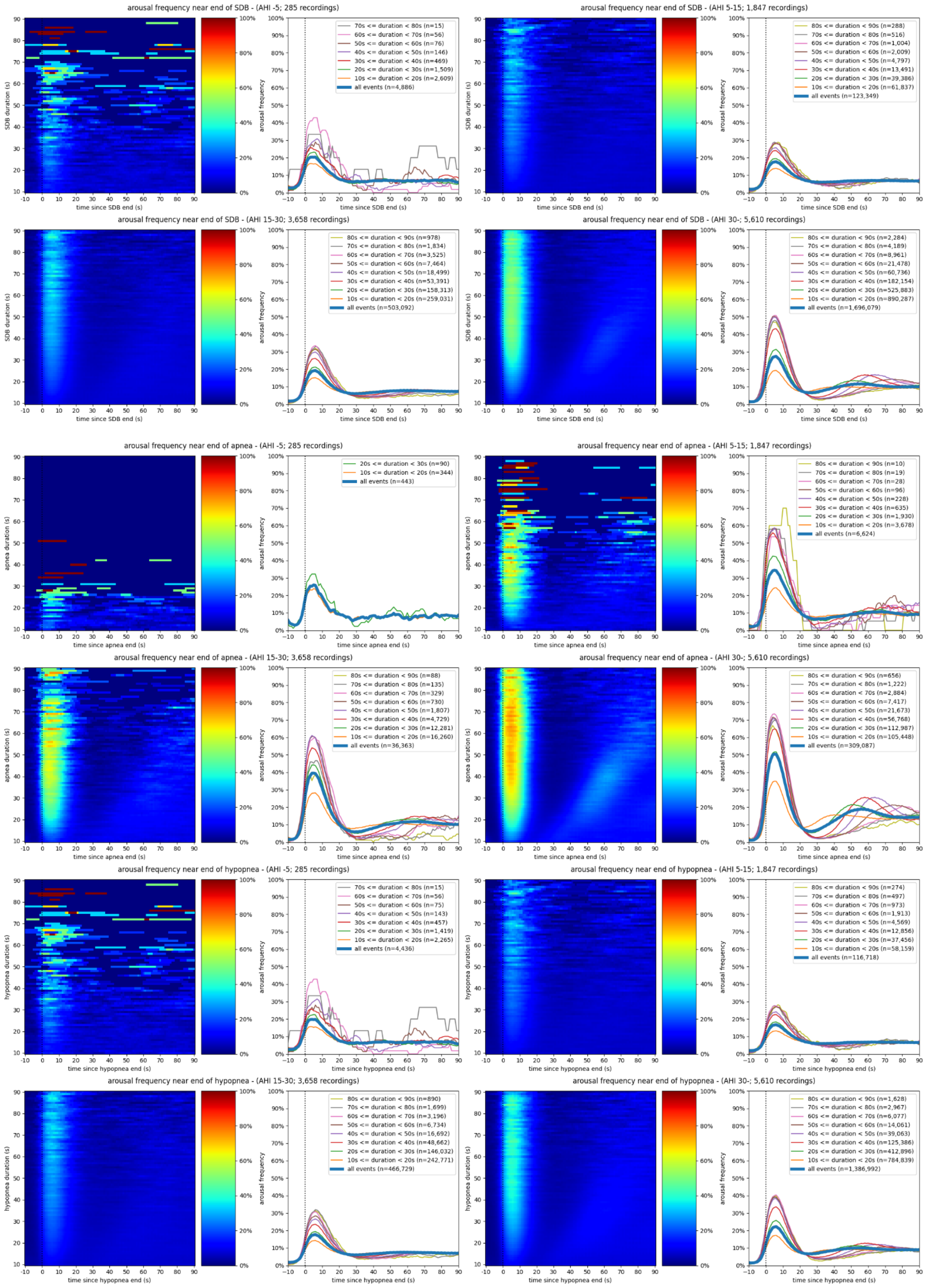
arousal frequency near end of SDB - (AHI -5; 285 recordings)
70s <= duration < 80s (n=15)
60s <= duration < 70s (n=56)
50s <= duration < 60s (n=76)
40s <= duration < 50s (n=146)
30s <= duration < 40s (n=469)
20s <= duration < 30s (n=1,509)
10s <= duration < 20s (n=2,609)
all events (n=4,886)
arousal frequency near end of SDB - (AHI 5-15; 1,847 recordings)
80s <= duration < 90s (n=288)
70s <= duration < 80s (n=516)
60s <= duration < 70s (n=1,004)
50s <= duration < 60s (n=2,009)
40s <= duration < 50s (n=4,797)
30s <= duration < 40s (n=13,491)
20s <= duration < 30s (n=39,386)
10s <= duration < 20s (n=61,837)
all events (n=123,349)
arousal frequency near end of SDB - (AHI 15-30; 3,658 recordings)
80s <= duration < 90s (n=978)
70s <= duration < 80s (n=1,834)
60s <= duration < 70s (n=3,525)
50s <= duration < 60s (n=7,464)
40s <= duration < 50s (n=18,499)
30s <= duration < 40s (n=53,391)
20s <= duration < 30s (n=158,313)
10s <= duration < 20s (n=259,031)
all events (n=503,092)
arousal frequency near end of SDB - (AHI 30-; 5,610 recordings)
80s <= duration < 90s (n=2,284)
70s <= duration < 80s (n=4,189)
60s <= duration < 70s (n=8,961)
50s <= duration < 60s (n=21,478)
40s <= duration < 50s (n=60,736)
30s <= duration < 40s (n=182,154)
20s <= duration < 30s (n=525,883)
10s <= duration < 20s (n=890,287)
all events (n=1,696,079)
SDB duration (s)
time since SDB end (s)
arousal frequency
arousal frequency near end of apnea - (AHI -5; 285 recordings)
20s <= duration < 30s (n=90)
10s <= duration < 20s (n=344)
all events (n=443)
arousal frequency near end of apnea - (AHI 5-15; 1,847 recordings)
80s <= duration < 90s (n=10)
70s <= duration < 80s (n=19)
60s <= duration < 70s (n=28)
50s <= duration < 60s (n=96)
40s <= duration < 50s (n=228)
30s <= duration < 40s (n=635)
20s <= duration < 30s (n=1,930)
10s <= duration < 20s (n=3,678)
all events (n=6,624)
arousal frequency near end of apnea - (AHI 15-30; 3,658 recordings)
80s <= duration < 90s (n=88)
70s <= duration < 80s (n=135)
60s <= duration < 70s (n=329)
50s <= duration < 60s (n=730)
40s <= duration < 50s (n=1,807)
30s <= duration < 40s (n=4,729)
20s <= duration < 30s (n=12,281)
10s <= duration < 20s (n=16,260)
all events (n=36,363)
arousal frequency near end of apnea - (AHI 30-; 5,610 recordings)
80s <= duration < 90s (n=656)
70s <= duration < 80s (n=1,222)
60s <= duration < 70s (n=2,884)
50s <= duration < 60s (n=7,417)
40s <= duration < 50s (n=21,673)
30s <= duration < 40s (n=56,768)
20s <= duration < 30s (n=112,987)
10s <= duration < 20s (n=105,448)
all events (n=309,087)
apnea duration (s)
time since apnea end (s)
arousal frequency near end of hypopnea - (AHI -5; 285 recordings)
70s <= duration < 80s (n=15)
60s <= duration < 70s (n=56)
50s <= duration < 60s (n=75)
40s <= duration < 50s (n=143)
30s <= duration < 40s (n=457)
20s <= duration < 30s (n=1,419)
10s <= duration < 20s (n=2,265)
all events (n=4,436)
arousal frequency near end of hypopnea - (AHI 5-15; 1,847 recordings)
80s <= duration < 90s (n=274)
70s <= duration < 80s (n=497)
60s <= duration < 70s (n=973)
50s <= duration < 60s (n=1,913)
40s <= duration < 50s (n=4,569)
30s <= duration < 40s (n=12,856)
20s <= duration < 30s (n=37,456)
10s <= duration < 20s (n=58,159)
all events (n=116,718)
arousal frequency near end of hypopnea - (AHI 15-30; 3,658 recordings)
80s <= duration < 90s (n=890)
70s <= duration < 80s (n=1,699)
60s <= duration < 70s (n=3,196)
50s <= duration < 60s (n=6,734)
40s <= duration < 50s (n=16,692)
30s <= duration < 40s (n=48,662)
20s <= duration < 30s (n=146,032)
10s <= duration < 20s (n=242,771)
all events (n=466,729)
arousal frequency near end of hypopnea - (AHI 30-; 5,610 recordings)
80s <= duration < 90s (n=1,628)
70s <= duration < 80s (n=2,967)
60s <= duration < 70s (n=6,077)
50s <= duration < 60s (n=14,061)
40s <= duration < 50s (n=39,063)
30s <= duration < 40s (n=125,386)
20s <= duration < 30s (n=412,896)
10s <= duration < 20s (n=784,839)
all events (n=1,386,992)
hypopnea duration (s)
time since hypopnea end (s)

Figure S2. Heatmap showing cortical arousal probability as a function of event duration and time since the end of the event for PSG recordings with AHI<5, AHI=5-15, AHI=15-30, and AHI>30 for SDB events (top four panels), for apnea events (middle four panels), and for hypopnea events (bottom four panels). X-axis indicates time since the end of the event and ranges from 10 seconds before to 90 seconds after the end of the event, with 0 corresponding to the end of the event. Y-axis shows event duration and ranges from 10 seconds to 90 seconds. Color on the heatmap indicates cortical arousal probability in 1-second resolution (i.e., likelihood of an arousal to be present at a given second) and ranges from 0% (dark blue) to 100% (dark red). Arousal probability curves next to the heatmaps are a quantification of the data in the heatmaps and indicate arousal probability (y-axis) for each second of the data (x-axis) for a given event duration (line color).
